# Interictal Epileptiform Discharge Detection Using Probabilistic Diffusion Models and AUPRC Maximization

**DOI:** 10.1101/2025.05.23.25328198

**Authors:** Lantian Zhang, Duong Nhu, Yun Zhao, Emma Foster, Lyn Millist, Shobi Sivathamboo, Patrick Kwan, Zongyuan Ge, Lan Du, Levin Kuhlmann

## Abstract

Recently, automated Interictal Epileptiform Discharge (IED) detection has attracted significant attention as a challenging predictive data analysis task aimed at improving early epilepsy diagnosis. Automated IED detection simplifies visual inspection and assists clinicians in identifying crucial IED waveform patterns in electroencephalographic (EEG) brain activities. However, IEDs are vastly outnumbered by non-IED or background data, directly training on such data leads to a detrimental impact on model performance. Moreover, most existing methods lack high precision when tested on cross-institution datasets and this will lead to time wasted by the neurologist having to look at predicted IEDs that are not IEDs. To address these issues, we propose a novel approach that employs probabilistic diffusion models for data augmentation alongside AUPRC maximization methods. This approach effectively addresses data scarcity and imbalance by combining real and synthesized IEDs to fully balance the training dataset, resulting in a 4.5% increase in precision and a 0.7% improvement in the F1-score during within-data evaluation. Additionally, it proves to be robust and generalizable, as evidenced by a 40.04% and 18.74% enhancement in precision and F1-score when applied to cross-data evaluation using data from another hospital.

## 1. Introduction

Neural signal analysis technologies involving AI and deep learning have been implemented to detect abnormal events such as interictal epileptiform discharges (IEDs), sleep disorders, and Alzheimer’s disease in the EEG (Siuly et al., 2024; Nhu et al., 2023; Aristimunha et al., 2023). However, the practical implementation of these technologies is hindered by issues related to the availability, imbalance, and low quality of EEG datasets. In this work, we focus on epilepsy diagnosis related IED detection tasks and employ diffusion probabilistic models as a data augmentation method to address these data issues. Additionally, we apply Average-Precision Loss (APLoss) as an auxiliary objective function to ensure precision.

IEDs are widely recognized as the most well-documented biomarkers of epilepsy (Kural et al., 2020). They are extensively used in clinical practice to confirm epilepsy, particularly in patients with ambiguous clinical presentations and those who have experienced their first seizure (Fisher et al., 2014). However, Identifying IEDs requires extensive training and specialized knowledge, often absent in resource-limited areas. AI models for automated IED detection show promise in bridging the gap and reducing the burden in centers with available expertise.

Over the past five decades, much research has been dedicated to automating the detection of IEDs. Early methods relied on predefined morphological and frequency features of EEG signals, mirroring visual feature extraction (Gotman and Gloor, 1976). Recently, ANN has revolutionized this field, with Convolutional Neural Networks (CNN) surpassing traditional IED classifiers in performance (Munia et al., 2023; Nhu et al., 2023). SpikeNet outperformed both expert interpretation and a commercial industry-standard IED detector in terms of calibration error (Jing et al., 2020). Another model was trained using the five fiducial landmarks of the waveform (onset, peak, trough, slow-wave, peak, offset) and generates output that is easily interpretable by human experts (Nascimento et al., 2023). A recent study introduced a convolutional neural network (HEAnet) trained to detect IEDs originating in the hippocampus, improving the sensitivity for diagnosing temporal lobe epilepsy from 50-58% to 63-67% without reducing specificity (Abou Jaoude et al., 2022). Despite the effectiveness of these approaches, they generally overlook two critical issues. First, non-IED data typically outnumber IED data. Training models directly on such imbalanced datasets leads to a bias towards negative samples, producing deceptively high specificity. Second, commonly used objective functions like Binary Cross-Entropy (BCE) tend to produce models with high sensitivity and specificity for this kind of data but do not always maximize precision, especially when tested on cross-hospital datasets (Nhu et al., 2023). Consequently, most works achieved promising results but still frequently give false positive predictions which will lead to wasted time by clinicians seeking to make diagnostic decisions based on IED predictions.

To address these data issues, one reliable approach is to use generative models to synthesize IEDs. Generative models such as GAN (Goodfellow et al., 2014), VAE (Kingma, 2013), and Diffusion models (Ho et al., 2020) have gained great success in image synthesis tasks. However, GANs often suffer from mode collapse due its instability and are prone to miss modes while training (Kushwaha et al., 2020), and VAEs tend to produce low-quality data due to its limited expressiveness in the latent space (Yang et al., 2024). Consequently, diffusion models tend to be a better option but have not yet been applied to IED data augmentation. Diffusion models can be categorized as probabilistic (Ho et al., 2020) and score-based diffusion models (Song et al., 2020b). Works (Song et al., 2023, 2020a) have accelerated the data generation process, albeit with a compromise in synthesized data quality. EEG synthesis employing DPMs can be broadly categorized as raw EEG synthesis (Torma and Szegletes, 2023) and latent EEG synthesis (Sharma et al., 2023). In this work, we apply a forward and reverse process (Ho et al., 2020) and use an architecture called EEGWave (Torma and Szegletes, 2023) to synthesize normalized single channel IED data. The synthesized IEDs are quantitatively evaluated for data quality, including fidelity and diversity. They are then combined with real IEDs to balance the training dataset, mitigating issues of data scarcity and imbalance.

To ensure precision in IED detection, one reliable approach is to directly optimize the Area Under the Precision-Recall Curve (AUPRC). AUPRC is a crucial metric in IED detection as it effectively demonstrates a model’s performance in reducing the false positive rate and maximizing precision. However, direct optimization of AUPRC encounters challenges related to its non-differentiable and non-decomposable nature. To address the nondifferentiable challenge, mainstream works have substituted non-differentiable loss functions with surrogate loss functions such as exponential loss (Qin et al., 2008), sigmoid loss (Brown et al., 2020), and linear interpolation function (Jiang et al., 2020). For the non-decomposable challenge, these works typically employ mini-batch methods to calculate average precision (Brown et al., 2020; Qin et al., 2010). Despite these efforts, none of these methods can guarantee convergence. Recently, Qi et al. (2021) proposed a compositional optimization method that provides a provable convergence guarantee. In this work, we use APLoss as defined by Qi et al. (2021).

The main contributions of this work are as follows. (1) We use diffusion modeling to synthesize normalized IEDs to address the scarcity, imbalance, and quality issues. (2) Through Data Augmentation with the auxiliary objective function, we separately enhance the precision by 4.5% and F1-score by 0.7% in within-data evaluation for a single hospital dataset. (3) Our approach is also applicable for cross-hospital data evaluation, as evidenced by a 40.04% and 18.74% increase in precision and F1-score on a dataset obtained from different hospitals.

## 2. Proposed Method

### 2.1. datasets

This study utilizes two datasets: the Temple University Events (TUEV) corpus and the Alfred Hospital datasets. The TUEV corpus, part of the Temple University Hospital (TUH) datasets, contains data from 390 patients, divided into distinct training and evaluation sets. This publicly available dataset employs a Temporal Central Parasaggital (TCP) montage, featuring 19 EEG electrodes and 20 reference channels placed on each patient’s scalp. At Temple University Hospital, the TUEV dataset contains six distinct events, as detailed in Table 1. The first three events (SPSW, GPED, and PLED) are clinically identified as interictal epileptiform discharges (IED) events, while the remaining three (EYEM, ARTF, and BCKG) are categorized as non-IED events. This classification allows for channel-wise binary classification on the TUEV corpus. Channel-wise classification refers to the detection of IEDs in individual EEG channels, as opposed to epoch-wise detection, which involves labeling IEDs across all EEG channels within the same time epoch.

**Table 1:** The number and distribution of events in the TUEV datasets—Training, Validation, and Evaluation—are used for channel-wise classification.

| Events |  | Channel-Wise |  |  |
| --- | --- | --- | --- | --- |
|  |  | Training Dataset<br>Count (%) | Validation Dataset<br>Count (%) | Evaluation Dataset<br>Count (%) |
| IED | Spike and/or sharp waves (SPSW) | 331<br>(0.46) | 312<br>(2.75) | 567<br>(1.93) |
|  | Generalized periodic epileptiform discharge (GPED) | 11106<br>(15.30) | 148<br>(1.31) | 4677<br>(15.90) |
|  | Periodic lateralized epileptiform discharge (PLED) | 4868<br>(6.71) | 1295<br>(11.43) | 1998<br>(6.79) |
| Non-IED | Eye movement (EYEM) | 692<br>(0.95) | 378<br>(3.34) | 329<br>(1.11) |
|  | Artifacts (ARTF) | 6428<br>(8.86) | 4625<br>(40.81) | 2204<br>(7.49) |
|  | Background (BKCG) | 49151<br>(67.72) | 4575<br>(40.37) | 19646<br>(66.78) |
| Total single channel IED segments |  | 16305<br>(22.47) | 1755<br>(15.49) | 7242<br>(24.61) |
| Total single channel non-IED segments |  | 56271<br>(77.53) | 9578<br>(84.51) | 22179<br>(75.38) |
| Total single channel segments |  | 72576<br>(100) | 11333<br>(100) | 29421<br>(100) |

The Alfred dataset, approved by the Alfred Health Ethics Committee (Project No: 745/19), was collected from two hospitals in Australia and comprises records of idiopathic generalized epilepsy (IGE) from 120 patients. The data is annotated in an epoch-wise manner, distinguishing between two classes: those containing interictal epileptiform discharges (IED) and those without. Each IED epoch begins one hundred to two hundred milliseconds before the first noticeable spike or rhythm change and concludes with the last slow wave before returning to the predischarge background. The dataset adheres to the same TCP montage utilized in the TUEV corpus. A detailed summary of this dataset is provided in Table 2.

**Table 2:**
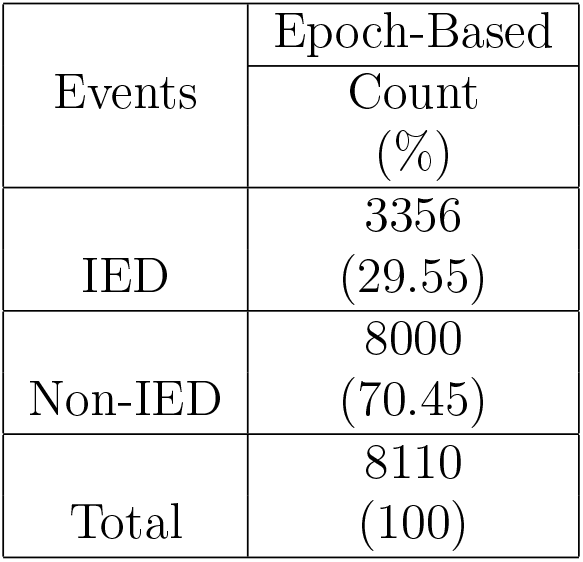
Number and distribution of events in Alfred Evaluation dataset.

### 2.2. Preprocessing and Transforming

To ensure the consistency between the two datasets, we removed electrodes M1 and M2 from the TCP montage as they are absent in some recordings of the Alfred dataset. Following this, we applied a 0.5-49Hz bandpass filter to remove low-frequency artifacts, high-frequency muscle artifacts, and power line noise. This filtering process aligns with established methodologies in the field (Nhu et al., 2023; Munia et al., 2023). Subsequently, we resampled the data to 256 Hz to obtain 1-second long single-channel segments from the TUEV dataset. For the Alfred dataset, we employed a sliding window with a 0.5-second step size to process IED epochs longer than 1 second, thereby extracting 1-second multi-channel epoch data. For non-IED epochs in the Alfred dataset, we randomly chose annotation files that do not contain any spike activities and employed a sliding step (40 seconds) and window size (1 second) to obtain a fixed number of non-IED epochs. For TUEV Corpus, 10% of data is used as the validation dataset to search for optimal threshold as demonstrated in Table 1, whereas all Alfred data is employed for evaluation as demonstrated in Table 2. Finally, we normalized the epoch-wise and channel-wise data using z-score normalization.

### 2.3. AUPRC Maximization

Area under precision-recall curve (AUPRC) is one of the most important metrics in IED classification, since AUPRC reflects a more comprehensive measure of precision and sensitivity (recall) across multiple prediction thresholds. Bamber (1975) describes AUPRC as the average precision calculated by the score of a defined threshold, as shown in Equation 1.

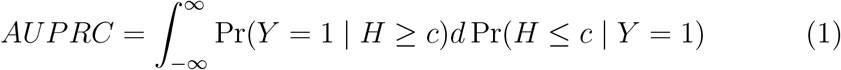

The precision at a given threshold *c* is represented by Pr(*Y* = 1| *H ≥ c*). In contrast, sensitivity is expressed as Pr(*H ≥ c* |*Y* = 1) = 1 − Pr(*H* ≤*c* |*Y* = 1). This equation represents the integral between precision and recall, with *c* being a randomly defined threshold between 0 and 1. In response to non-differentiability issues, AUPRC can be approximated using Equation 2 as discussed in Qi et al. (2021).

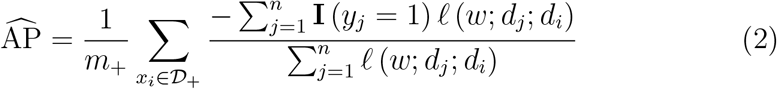

In this context, *m*_+_ and *𝒟*_+_ represent the number of positive samples and the positive sample set, respectively, while *n* denotes the total number of datasets. The surrogate loss functions, denoted as *ℓ*(*w*; *d*_*j*_; *d*_*i*_), are pivotal in this analysis (Qin et al., 2008; Brown et al., 2020; Jiang et al., 2020). Among these, Qi et al. (2021) introduced the squared hinge loss function *ℓ* (*w*; *d*_*j*_, *d*_*i*_) = max (0, *m* − (*g*_**w**_ (*d*_*i*_) − *g*_**w**_ (*d*_*j*_)))^2^ and the sigmoid loss function 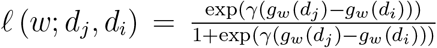. Here, *g* (*d*) represents the prediction score derived from neural networks of a sample *d*_*i*_, with *m* and *γ* serving as the margin and scaling factors, respectively. In our study, we employ the squared hinge loss function proposed by Qi et al. (2021) due to its demonstrated convergence properties.

### 2.4. Diffusion Model and Data Augmentation

Diffusion Probabilistic Models (DPM), initially introduced by Sohl-Dickstein et al. (2015), have demonstrated significant success in image and audio synthesis tasks (Rombach et al., 2022; Chen et al., 2020). These models are based on the principles of a Markov chain and encompass two primary processes: the forward diffusion process and the reverse sampling process. In the forward process, a sample *x*_0_ from the real data distribution is gradually transformed into a sample *x*_*T*_ from a simpler distribution, such as a Gaussian distribution, through the systematic addition of noise at each step. Conversely, the reverse process involves iteratively denoising the sample *x*_*T*_ ~*𝒩* (0, **I**) at each time step, thereby progressively reconstructing a sample that approximates the original data distribution. The overview of Diffusion and Reverse Markov chain is shown in Figure 1.

**Figure 1:**
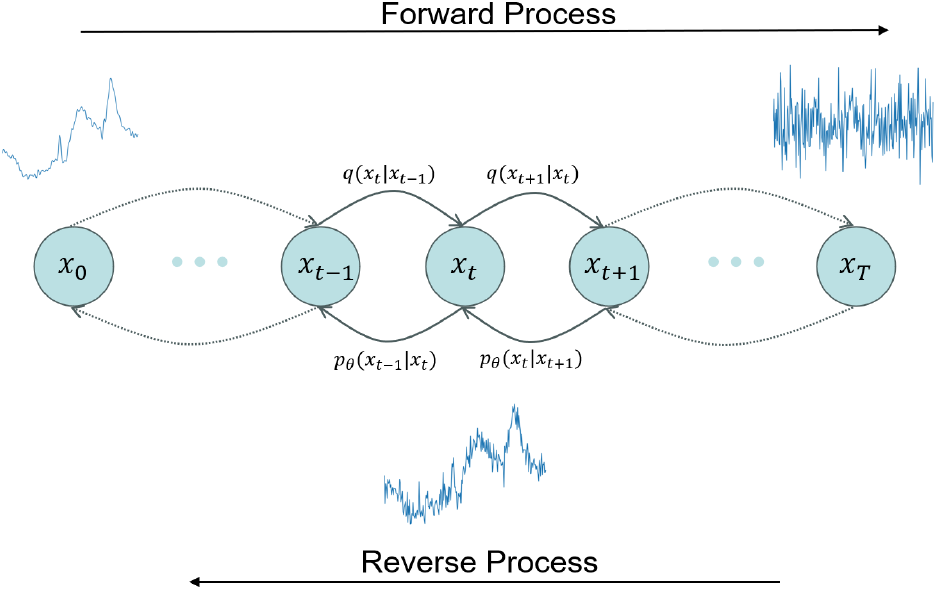
Overview of forward and reverse Markov chain

The objective of DPMs is to maximize the variational lower bound. By using parameterization and Langevin dynamic ensembling as described in the work (Ho et al., 2020), the proposed objective function can be simplified to a simple L1 or L2 loss function. The model architecture we used is called EEGWave proposed in work (Torma and Szegletes, 2023). The overall architecture is demonstrated in Fig 2.

**Figure 2:**
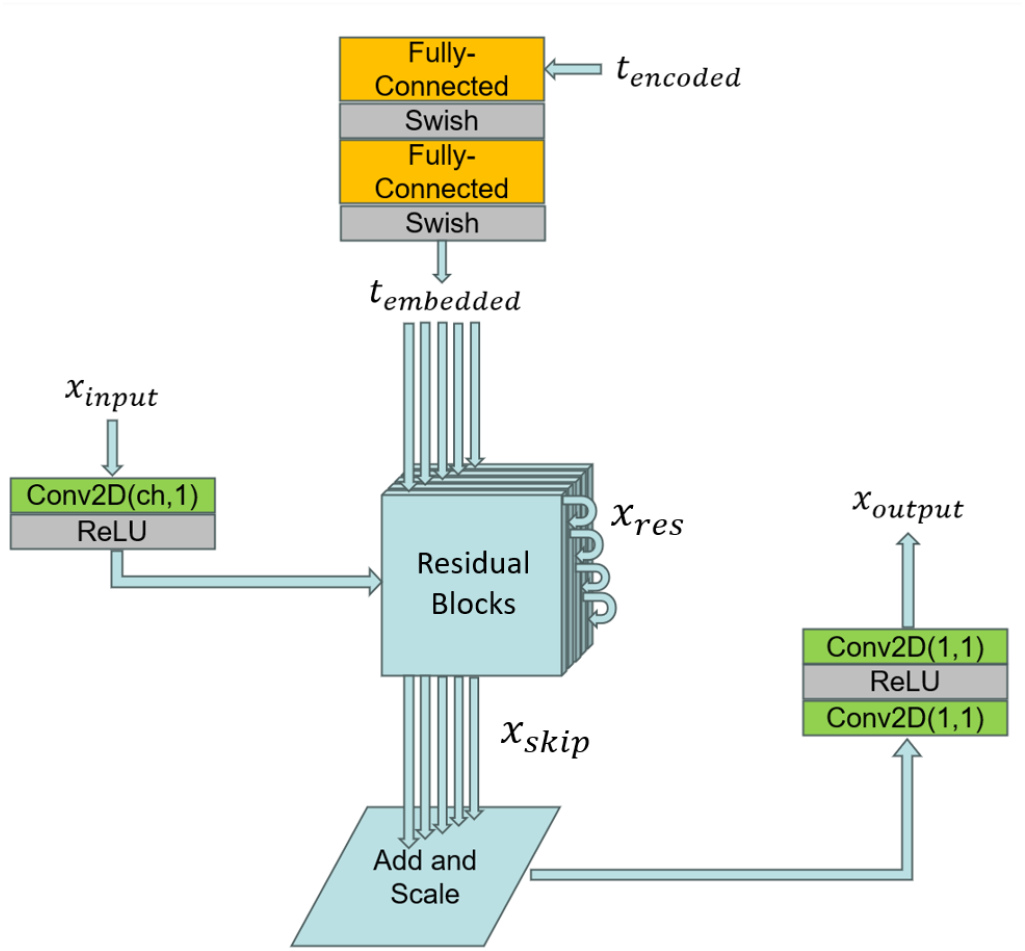
Architecture of EEGWave

EEGWave is a novel diffusion model designed as a sequence-to-sequence model with identical input and output dimensions. Initially, the perturbed data passes through a spatial convolution layer and an activation function to capture cross-channel features. Then the extracted features are passed to the first layer of the res-block.

Another input to the res-block layers is time-embedding. We employ positional embedding to encode a single-time step into a 128-length vector, as depicted in the work (Vaswani, 2017). Following that, two fully connected layers using the Swish activation function are applied to pass the time step embedding to the res-block.

The model contains a group of res-blocks, where encoded perturbed data and time step embedding are added to the temporal convolution, enabling the residual block to learn local characteristics. Following this, the result is split along channels into two parts. Sigmoid and Tanh activation are separately applied to each part and element-wise multiplication is applied. After this, point-wise convolution is used and the data is again split along channels into two chunks. One chunk is a direct output and the other is the input to the next res-block.

After deriving the direct output from each res-block, the outputs are summed and normalized by the number of res-blocks. Then, they pass through two 1×1 convolution kernels with ReLU as the activation function to obtain the desired output channel.

In this work, we use 40 residual blocks with a dilation cycle of 2^0:7^. The input channel and output channel are 1 and we set the kernel size equal to 3. The perturbed input embedding and time step embedding are both 512 dimensional such that they can do element-wise addition. We employ a 1000-step linear schedule, with *β*_*t*_ ranging from 1 × 10^−4^ to 0.02. The diffusion and reverse diffusion process of this work follows algorithms proposed in works (Ho et al., 2020; Torma and Szegletes, 2023).

### 2.5. Classifiers

#### 2.5.1. InceptionTime

The Inception architecture, introduced by Szegedy et al. (2016, 2017), employs multiple kernel sizes and pooling operation in parallel to derive features at different scales. Inspired by this, InceptionTime (Ismail Fawaz et al., 2020) employed 1D convolution with different kernel sizes to extract both short and long term feature and achieved SOTA result on the UCR archive. We use code from the tsai library (Oguiza, 2023), where they provide SOTA techniques for time series tasks such as classification, regression, and imputation. The Inception Module is demonstrated in Fig 3 and the whole architecture is demonstrated in Fig 4.

**Figure 3:**
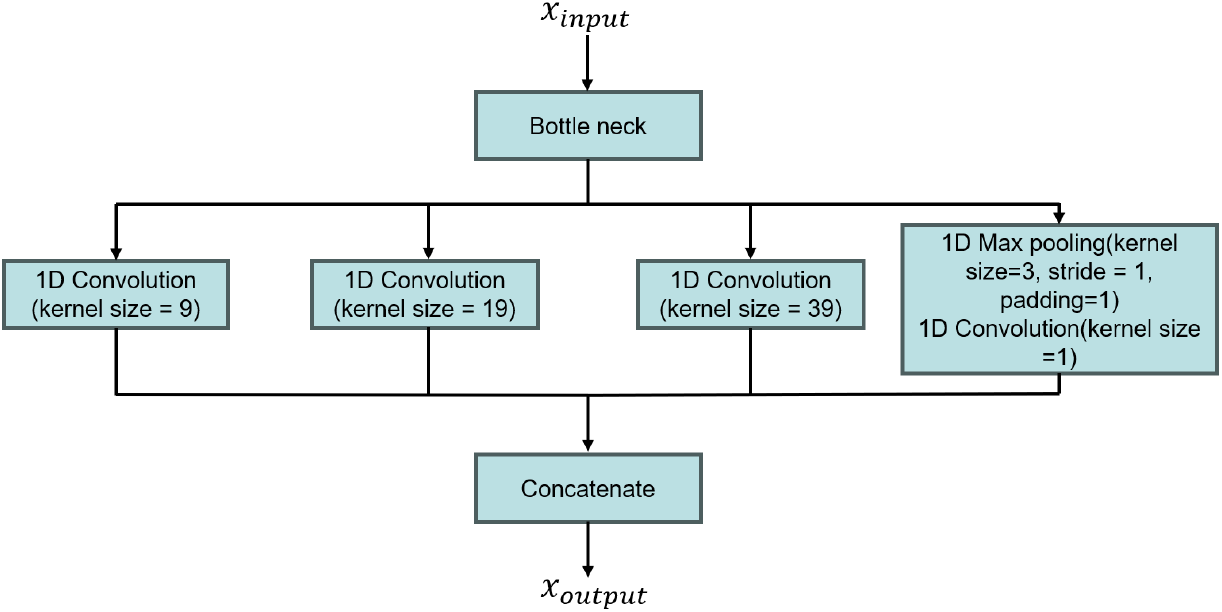
Architecture of Inception Module

**Figure 4:**
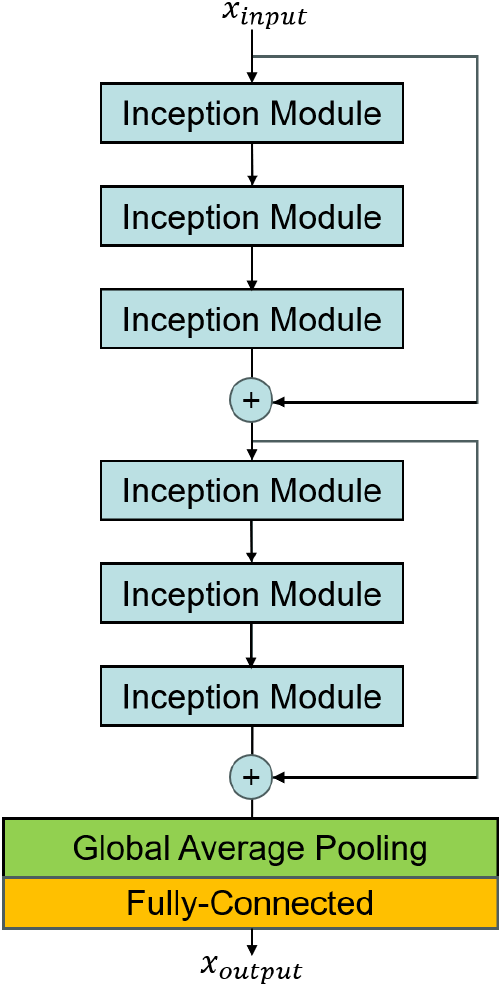
Architecture of InceptionTime

#### 2.5.2. Minirocket

Another Classifier we use is Minirocket (Dempster et al., 2021). It involves feature Engineering with a fixed set of kernels to transform time series data. The transformed data is then used to train a linear classifier. Through this approach, it greatly reduces the computation time while maintaining performance.

#### 2.5.3. S4D-ECG

One more classifier we use is S4D-ECG (Huang et al., 2024), where they integrate the Diagonal State Space Sequence (S4D) model to capture the inner dynamics of ECG data. Through this model architecture, they achieved competitive results with explainable model architecture.

### 2.6. Overall Architecture

The overall architecture of this work, depicted in Fig 5, comprises three main steps. First, a Denoising Probabilistic Model (DPM) is trained using perturbed data *x*_*t*_. Second, noise is iteratively subtracted from samples drawn from a simple Gaussian distribution, allowing us to derive synthesized IEDs. Finally, these synthesized IEDs are combined with normalized IEDs and input into time series classifiers for training.

**Figure 5:**
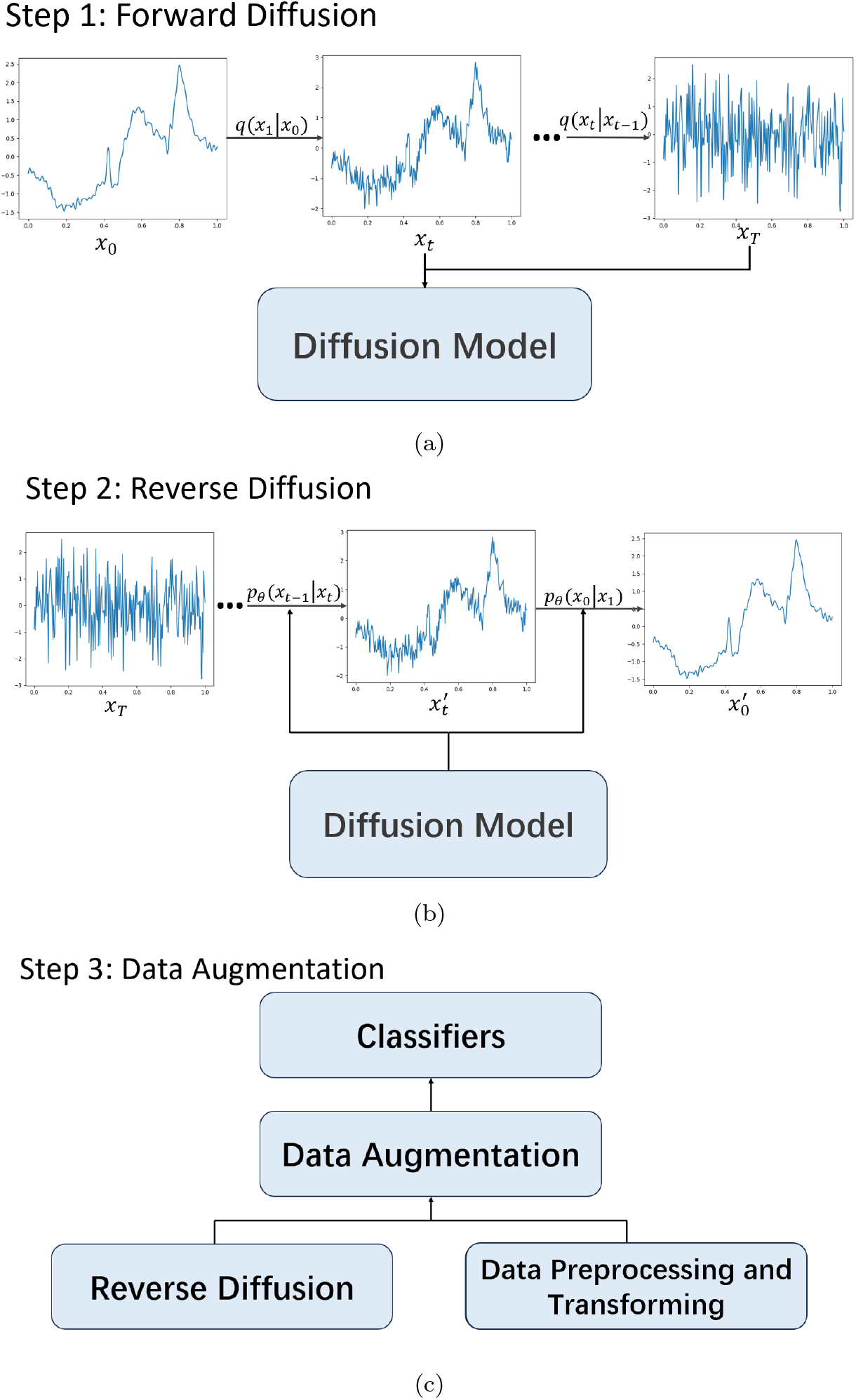
The Overall architecture of our approach. (a) Training denoising probabilistic diffusion model with perturbed *x*_*t*_. (b) Reverse diffusion to synthesize normalized single channel IED data. (c) Combine synthesized IEDs with real IEDs through data augmentation and then pass them into classifiers.

### 2.7. Metrics

#### 2.7.1. Metrics for classifiers

In this work, we use AUC, AUPRC, sensitivity, precision, and F1-score as metrics for time series classifiers. Sensitivity demonstrates the percentage of annotated IEDs correctly identified as IEDs, while precision demonstrates the proportion of identified IEDs that are annotated IEDs. An effective algorithm must balance precision and sensitivity. As such, we use F1-score, capturing the balance and demonstrating the overall performance of both sensitivity and precision. In addition to those metrics, AUC and AUPRC are also used to evaluate the performance of the classifiers.

#### 2.7.2. Metrics for synthesized IEDs

In this section, we employ Fréchet Inception Distance (FID), density, and coverage to assess the fidelity and diversity of synthesized IEDs. FID, introduced by Heusel et al. (2017), is the most widely and commonly employed metric in generative modeling with its formulation defined as follows:

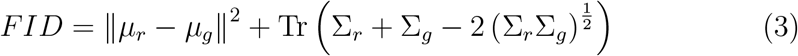

where *µ*_*r*_ and *µ*_*g*_ are the mean vectors of real and synthesized embeddings and Σ_*r*_ and Σ_*g*_ the correlation matrix of real and synthesized embeddings. In this work, the real and synthesized embeddings are generated from a pre-trained InceptionTime classifier without global average pooling and fully-connected layers. However, FID is under the strong assumption that the real and fake embeddings are drawn from a multivariate normal distribution, and a pre-trained classifier is commonly used to extract features of input data.

As such, we also use density and coverage metrics introduced by Naeem et al. (2020) to measure the fidelity and variety of synthesized data. The definition is shown as:

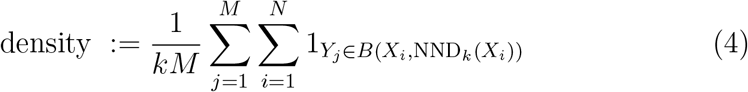

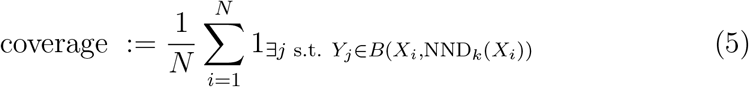

where *X*_*i*_ and *Y*_*j*_ are real and fake samples. *N* and *M* are the number of fake and real samples. *B* (*X*_*i*_, NND_*k*_ (*X*_*i*_)) is the sphere in ℝ^*D*^ around point *X*_*i*_ with radius equal to the distance NND_*k*_ (*X*_*i*_) between *X*_*i*_ and its *k*^th^ neighbor. This calculation avoids bias from pretrained models and strong assumptions by directly comparing synthesized and real data.

## 3. RESULTS

In this section, we present the quantitative result of synthesized IEDs. We then demonstrate the performance of our approaches for within-hospital data evaluation and subsequently evaluate their performance on cross-hospital data evaluation.

Table 3 presents the quantitative result of synthesized data through three metrics: FID, density, and diversity. To ensure a fair comparison between real and synthesized data, we individually trained an unconditioned DPM for each type of data. We synthesized the same number of data as demonstrated in Table 1. The metrics for PLED, GPED, and ARTF indicate good fidelity and variety, while the metrics of SPSW and EYEM are hindered due to limited training samples. Furthermore, we excluded BCKG as it lacks neurological relevance.

**Table 3:** Metrics of IEDs and non-IEDs.

| Data Type |  | Number | FID | Density | Coverage |
| --- | --- | --- | --- | --- | --- |
| IED | SPSW | 645 | 2.42 | 0.16 | 0.33 |
|  | PLED | 5965 | 0.10 | 1.07 | 0.91 |
|  | GPED | 11254 | 0.21 | 1.06 | 0.78 |
| non-IED | ARTF | 11053 | 0.17 | 1.10 | 0.87 |
|  | EYEM | 1070 | 1.77 | 0.30 | 0.36 |

### 3.1. Within-hospital data Evaluation

In this section, we train our models on the TUEV corpus training dataset, select threshold with highest F1-score based on validation dataset, and use evaluation dataset in Table 1 to test performance of different approaches. The performance of different approaches is illustrated in Table 4. Through data Augmentation, we achieved the highest AUPRC and F1-score, while through a combination of data augmentation and APLoss, we achieved the highest precision.

**Table 4:** Benchmarking of different approaches in within-hospital (TUEV) data evaluation.

| Method | AUC | AUPRC | Sens | Prec | F1 |
| --- | --- | --- | --- | --- | --- |
| Minirocket | $0.9891 \pm 0.0003$ | $0.9721 \pm 0.0005$ | **$0.9455 \pm 0.0031$** | $0.8724 \pm 0.0044$ | $0.9075 \pm 0.0014$ |
| S4D-ECG | **$0.9917 \pm 0.0005$** | $0.9792 \pm 0.0007$ | $0.9248 \pm 0.0072$ | $0.9316 \pm 0.0089$ | $0.9280 \pm 0.0013$ |
| InceptionTime | $0.9914 \pm 0.0007$ | $0.9788 \pm 0.0015$ | $0.9443 \pm 0.0128$ | $0.9067 \pm 0.0135$ | $0.9234 \pm 0.0041$ |
| InceptionTime† | $0.9917 \pm 0.0010$ | **$0.9800 \pm 0.0014$** | $0.9298 \pm 0.0047$ | $0.9309 \pm 0.0033$ | **$0.9303 \pm 0.0023$** |
| InceptionTime†† | $0.9911 \pm 0.0004$ | $0.9788 \pm 0.0008$ | $0.9081 \pm 0.0072$ | **$0.9477 \pm 0.0067$** | $0.9274 \pm 0.0014$ |

† After Data Augmentation
†† After Data Augmentation and use AP Loss as objective function

#### 3.1.1. The Effect of Data Augmentation

In this section, we check the performance of data augmentation by gradually increasing the number of IEDs until the training dataset is balanced. The result is illustrated in Fig 6. We use paired t-tests to evaluate the statistical significance between groups with subsequent Bonferroni correction for multiple comparisons. The precision and F1-score are statistically higher than that of before augmentation. Also, we notice a decreasing trend in sensitivity when increasing the imbalanced data ratio (IR); However, It does not present a statistically significant result.

**Figure 6:**
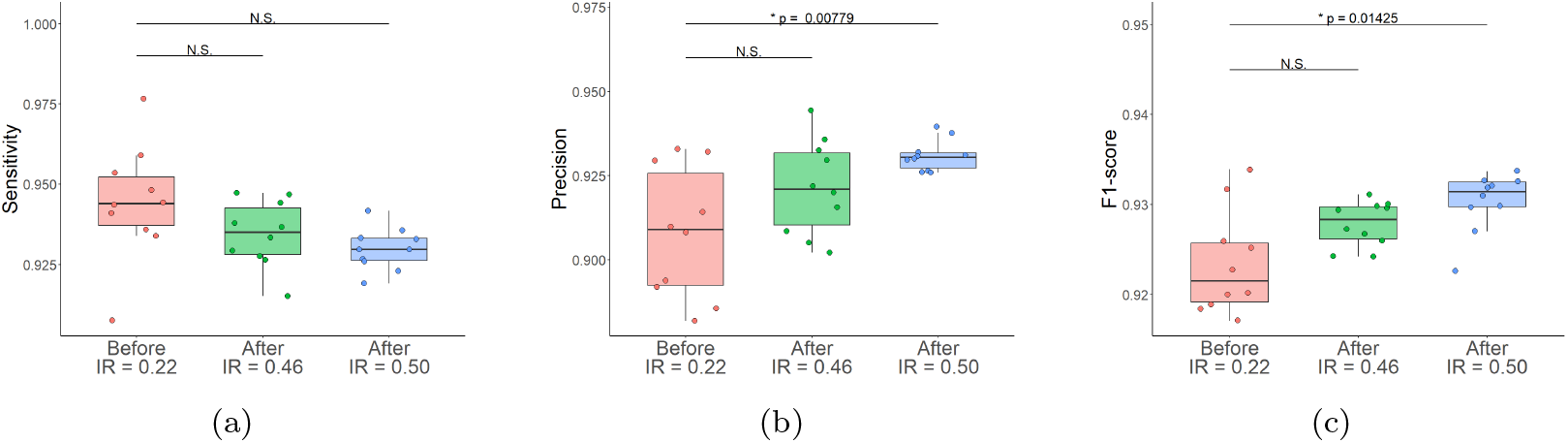
Effects of data augmentation for three imbalance ratios (IR) and within-hospital (TUEV) data evaluation. (a) Sensitivity, (b) Precision, and (c) F1-score of before (red: IR=0.22) and after (green: IR=0.46 and blue: IR = 0.5) data augmentation.

#### 3.1.2. The Effect of Objective Function

In this section, we test the performance of the objective function for different data imbalanced ratios. The result of sensitivity is shown in Fig 7. Similarly, using APLoss as an objective function achieves a trade-off between sensitivity and precision. The precision of APLoss tends to be higher than that of BCE, as APLoss directly optimizes precision. Also, when data is imbalanced with IR equal to 0.22, the F1-score tends to be higher as the definition of APLoss is more suitable for an imbalanced dataset.

**Figure 7:**
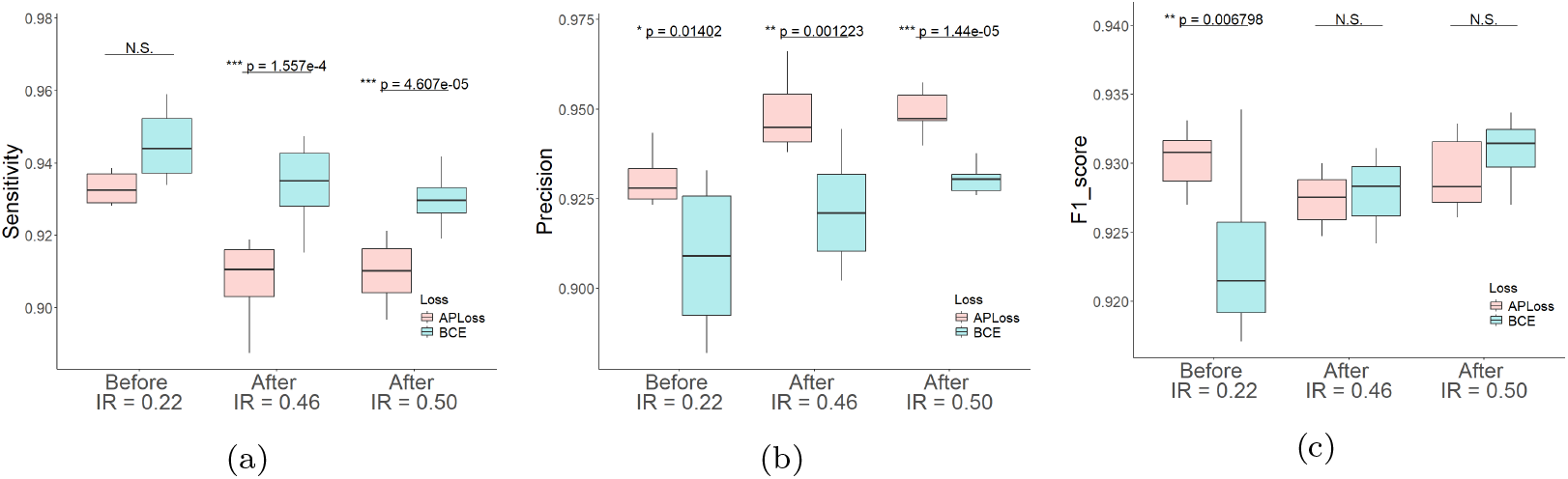
Performance metrics for different objective functions in within-hospital (TUEV) evaluation. (a) Sensitivity, (b) Precision, and (c) F1-score of APLoss (pink) and BCE Loss (blue).

### 3.2. Cross-Hospital Data Evaluation

In this section, we train our model on the TUEV corpus and evaluate its performance on the Alfred Hospital data, as presented in Table 2. We employ ensembling techniques across EEG channels, calculating the maximum probability for channels as the epoch probability. We then employ optimal threshold that maximizes the F1-score of TUEV validation set. This optimal threshold is subsequently applied to the Alfred evaluation dataset, and the evaluation outcomes are detailed in Table 5.

**Table 5:**
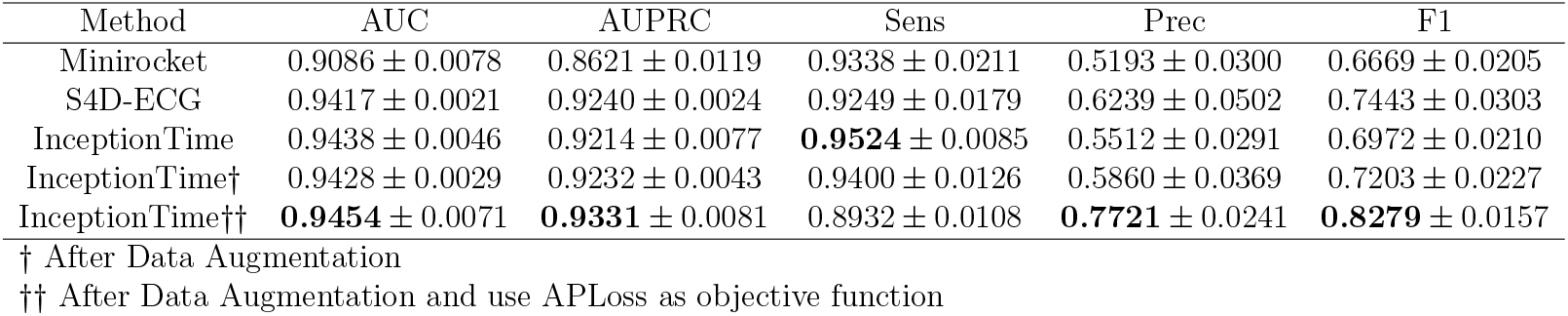
Benchmark of different approaches in cross-hospital (Alfred) evaluation.

| Method | AUC | AUPRC | Sens | Prec | F1 |
| --- | --- | --- | --- | --- | --- |
| Minirocket | 0.9086 ± 0.0078 | 0.8621 ± 0.0119 | 0.9338 ± 0.0211 | 0.5193 ± 0.0300 | 0.6669 ± 0.0205 |
| S4D-ECG | 0.9417 ± 0.0021 | 0.9240 ± 0.0024 | 0.9249 ± 0.0179 | 0.6239 ± 0.0502 | 0.7443 ± 0.0303 |
| InceptionTime | 0.9438 ± 0.0046 | 0.9214 ± 0.0077 | **0.9524** ± 0.0085 | 0.5512 ± 0.0291 | 0.6972 ± 0.0210 |
| InceptionTime† | 0.9428 ± 0.0029 | 0.9232 ± 0.0043 | 0.9400 ± 0.0126 | 0.5860 ± 0.0369 | 0.7203 ± 0.0227 |
| InceptionTime†† | **0.9454** ± 0.0071 | **0.9331** ± 0.0081 | 0.8932 ± 0.0108 | **0.7721** ± 0.0241 | **0.8279** ± 0.0157 |

† After Data Augmentation †† After Data Augmentation and use AP Loss as objective function

Through our approaches, we demonstrate that employing data augmentation alongside AUPRC maximization methods enhances precision and F1-score, as evidenced by analysis in Fig 8 with statistical significant results.

**Figure 8:**
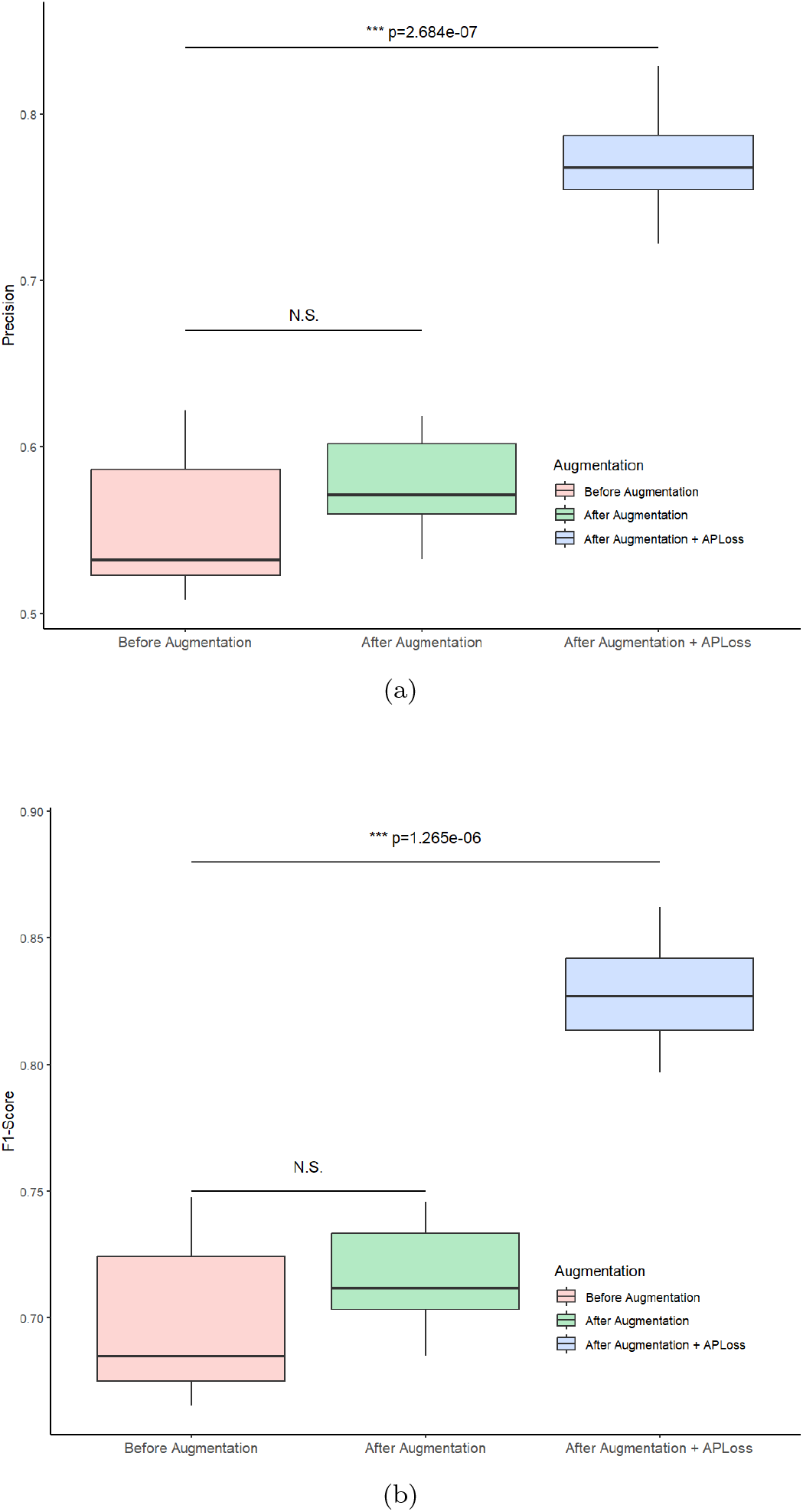
(a) Precision and (b) F1-score of cross-hospital (Alfred) data evaluation based on models trained in three cases (before augmentation, after augmentation, and after augmentation with APLoss).

## 4. Discussion

Mainstream works of IED detection (Nhu et al., 2023; Munia et al., 2023; Jing et al., 2020) have achieved promising results in IED detection. However, many fail to provide compelling evidence regarding its accuracy and clinical utility. Few papers include performance-metrics derived from independent test datasets, and even when they do, these datasets often lack the necessary size to adequately represent the wide variability in IEDs (Munia et al., 2023; Jing et al., 2020). Additionally, performance is evaluated on short epochs (1-2 seconds) and per-patient performance is frequently based on selected epochs (13-30 seconds) rather than full-length recordings (Nhu et al., 2023). The performance of IED-detectors assessed at individual level is both impossible and of questionable clinical relevance. EEG-fMRI studies have demonstrated that IEDs are often obscured by background activities (Grouiller et al., 2011), which means the true number of IEDs in scalp EEG recordings cannot be reliably determined. This inherent limitation cannot be overcome by relying on a majority expert consensus for IED classification. Moreover, the absolute number of IEDs typically is not part of clinical assessment. In a clinical setting, the critical question is whether the patient’s EEG contains IEDs or not. Therefore, reporting clinically relevant performance requires evaluation at the level of the entire EEG rather than the level of single discharges. Another significant challenge in conducting clinical validation studies of IED-detectors is establishing the reference standard, also known as the gold standard. Given that interrater reliability in identifying IEDs is often low, the gold standard serves as a benchmark to ensure accuracy and consistency. This standard can be determined in one of two ways: either through a majority consensus among a large group of experts, typically comprising 5 to 10 specialists (Halford et al., 2017) or ideally by employing an external diagnostic reference standard such as video-EEG recordings of habitual paroxysmal episodes or long-term follow-up.

Table 6 presents a comparative analysis of our study with other research efforts. In terms of sensitivity, precision, and F1-score, our work demonstrates promising results when compared to the studies conducted by Nhu et al. (2023); Munia et al. (2023). Additionally, we evaluated the AUC as indicated in works (Jing et al., 2020; Nascimento et al., 2023), aligning our metrics with those utilizing AUC and calibration error as evaluation criteria. Our findings indicate that we also achieved competitive results in the context of AUC as well.

**Table 6:** Metrics in various works. † after augmentation †† after augmentation and APLoss as objective function.

| Works | Dataset | AUC | Sensitivity | Precision | F1-score |
| --- | --- | --- | --- | --- | --- |
| (Munia et al., 2023) | TUEV Corpus | - | 0.86 | 0.88 | 0.87 |
| (Nhu et al., 2023) | TUEV Corpus | 0.99 | 0.90 | 0.93 | 0.92 |
| (Jing et al., 2020) | 1051 EEGs | 0.97 | - | - | - |
| (Nascimento et al., 2023) | 1051 EEGs | 0.88 | - | - | - |
| Ours | TUEV Corpus | 0.99 | 0.94 | 0.91 | 0.92 |
| Ours<sup>†</sup> | TUEV Corpus | 0.99 | 0.93 | 0.93 | 0.93 |
| Ours<sup>††</sup> | TUEV Corpus | 0.99 | 0.91 | 0.95 | 0.93 |

As described above, several AI-based IED detectors have achieved high sensitivity but at the cost of false detection. In the semi-automated approach, experts review the IED candidates identified by AI models to either accept or reject them. However, this process becomes time-consuming when the AI model generates a large number of candidates. A potential solution involves automatically grouping spikes with similar morphological and spatial characteristics into “clusters”. Experts then review a few examples from each cluster to make decisions about the entire group (Kural et al., 2020). This semi-automated approach enhances the specificity of IED detection. A similar approach was employed for long-term ambulatory EEGs, which reduced the time spent on visual analysis in the clinic by 50–75 times without compromising accuracy (da Silva Lourenço et al., 2023). Further research and development should focus on establishing large, multicenter, open datasets with annotated IEDs, along with the adoption of standardized methods for testing and reporting the performance of the detectors.

Diffusion models have achieved great success in previous work (Ho et al., 2020). These models are primarily categorized into two types: score-based diffusion models and probabilistic diffusion models. In our research, we employ probabilistic diffusion models due to their higher density and diversity compared to score-based models in this work. For similar reasons, we opt for an unconditional diffusion model rather than a conditional one. In this study, we employ Euclidean distance to calculate the similarity between samples. Although dynamic time warping (DTW) might better account for spike lags, it remains computationally intensive, even with downsampled time resolution. Future work will focus on enhancing the model architecture, stochastic processes, and guidance strategies to further improve the density and diversity of synthesized samples. This enhancement will enable us to better understand the relationship between classifier metrics and metrics of generative models.

We employ APLoss (Qi et al., 2021) as the objective function to achieve a trade-off between sensitivity and precision. According to the definition in Equation 2, APLoss is particularly suitable for imbalanced datasets. Consequently, its application in balanced data scenarios may not lead to improvements in the F1-score. This is evident in Fig 7c, where an increase in the number of IEDs results in a decrease in the F1-score; However, APLoss remains a viable option for ensuring precision, even in balanced data cases. In addition, threshold searching based on highest F1-score is significantly influenced by the balance of the validation dataset, and using APLoss as the objective function mitigates the effects of this imbalance. This may explain the noticeable improvement in precision and F1-score in Figure 8.

## 5. Conclusion

Automated IED detection has been a growing topic of interest in neural signal analysis; However, most works achieved promising results but still frequently give false positive results especially on unseen cross-hospital evaluation data. These false positives result in more inspection time by the clinician. In this work, we present a novel method using data augmentation based on probabilistic diffusion modeling and AUPRC maximization to ensure precision. The work illustrates good generalization in both within-hospital data evaluation and cross-hospital data evaluation in terms of minimizing false positives. This enables a more reliable algorithm for both clinicians and patients, such that patients can be diagnosed with greater efficiency and as a result receive more appropriate treatments in a timely fashion.

## Data Availability

The first dataset (TUEV) dataset is avaliable online at https://isip.piconepress.com/projects/tuh_eeg/. The seocnd Alfred dataset produced in the present study are available upon reasonable request to the authors

## Notes

### Competing Interest Statement

The authors have declared no competing interest.

### Funding Statement

This study did not receive any funding

### Author Declarations

Ethics committee/IRB of Alfred Health Ethics Committee gave ethical approval for this work

### Summary of Updates

author list updated in the both meta data and in the manuscript;

